# Generalizing intensive care AI across time scales in resource-limited settings

**DOI:** 10.64898/2026.04.23.26351588

**Authors:** Akshaya Devadiga, Pradeep Singh, Jhuma Sankar, Rakesh Lodha, Tavpritesh Sethi

## Abstract

Generalization remains a major barrier to the clinical deployment of artificial intelligence in critical care. Models developed using high-frequency physiological monitoring often experience performance degradation when applied across hospitals, patient populations, and monitoring environments that differ from those used during training. Although variability in temporal resolution is widespread in critical care, its impact on representation learning and model transferability remains poorly understood.

Here, we introduce the resolution-transfer task for physiological time series and present PhysioStack, a self-supervised physiological representation learning framework designed to evaluate whether representations learned from high-frequency monitoring data remain robust across clinically relevant monitoring frequencies without retraining. We introduce SafeICU, a longitudinal pediatric intensive care dataset spanning ten years of routinely collected clinical and physiological data from a tertiary-care hospital in India, including high-resolution vital signs recorded at 15-second intervals. Transformer-based masked language models were trained on 144,271 patient-hours of high-resolution physiological signals from 984 pediatric ICU stays to learn representations of heart rate, respiratory rate, oxygen saturation, and arterial blood pressure.

Representations learned at high temporal resolution demonstrated strong transferability across lower monitoring frequencies, consistently outperforming models trained directly on temporally aggregated data. Importantly, these representations generalized across patient populations, maintaining performance when evaluated on independent adult intensive care cohorts derived from the MIMIC-III and eICU databases without retraining. In a downstream early shock prediction task, PhysioStack achieved AUROC values of 0.82-0.86 and AUPRC values of 0.91-0.94 across 5-60-minute monitoring frequencies without retraining. Representations trained on pediatric data generalized to independent adult ICU cohorts, achieving AUROC 0.91-0.95 and AUPRC 0.94-0.97 without cohort-specific fine-tuning.

These findings demonstrate that PhysioStack learns transferable physiological representations that generalize across temporal resolutions and patient cohorts, supporting robust AI for critical care. To support further research, we publicly release the SafeICU database, comprising longitudinal vital signs, laboratory measurements, treatment records, microbiology, and admission and discharge, together with pretrained physiological representation models.

**Research in context:** *Evidence before this study:* Before undertaking this study, we searched PubMed, Web of Science, and Google Scholar for articles published between 2010, and 2026. Search terms included combinations of (“self-supervised learning” OR transformer OR “masked language model”) AND (“critical care” OR ICU); (“MIMIC” OR “eICU” OR EHR OR “multimodal data”) AND (“critical care” OR ICU); and (“physiological time series” OR “vital signs” OR monitoring) AND (generalization OR “temporal resolution” OR “sampling frequency”). The equivalent PubMed searches yielded 302, 6468, and 214561 records, respectively, reflecting the rapid growth of research in artificial intelligence, critical care datasets, and physiological monitoring. Manual screening of relevant reviews and original articles showed that existing studies primarily investigate self-supervised learning for downstream clinical prediction or evaluate generalizability across institutions and patient cohorts. However, nearly all studies develop and evaluate models using a single temporal resolution, treating monitoring frequency as fixed.

*Added value of this study:* This study introduces the concept of temporal resolution transfer for physiological representation learning, SafeICU, a large pediatric intensive care database, and PhysioStack, a self-supervised framework pretrained using high-resolution physiological signals trained and a generalizable model for early prediction of shock in low resource settings, validated in three data cohorts spanning 200+ ICUs. We demonstrate that representations learned from high-frequency monitoring remain robust across clinically relevant monitoring frequencies without additional pretraining and generalize across independent pediatric and adult intensive care cohorts. We further show that these transferable representations support downstream early shock prediction across temporal resolutions and publicly release both the SafeICU database and pretrained PhysioStack models to support reproducible research and external benchmarking.

*Implications of all the available evidence:* Current evidence suggests that self-supervised learning can produce reusable representations for clinical prediction, but deployment remains limited by poor generalizability across heterogeneous healthcare environments. Our findings indicate that pretrained physiological representations can be transferred across differences in monitoring frequency while maintaining downstream predictive performance, reducing the need to retrain models for each sampling resolution. Together with existing evidence, these results support the development of more scalable and deployable physiological AI systems and provide a publicly available pediatric resource to accelerate future research on representation learning, external validation, and foundation models for critical care.

## 1. Main

Continuous physiological monitoring forms the foundation of modern intensive care by providing real-time assessment of a patient’s evolving clinical condition. In critically ill children, changes in vital signs frequently precede overt clinical deterioration, enabling earlier recognition of life-threatening conditions such as shock, sepsis, and respiratory failure when interpreted in the context of continuously acquired physiological data [1,2]. Consequently, longitudinal physiological time-series have become a central source of information for bedside decision-making and the development of predictive models that support early risk stratification and timely clinical intervention [1–4]. As monitoring technologies become increasingly integrated into routine critical care, the ability to learn robust representations from high-frequency physiological data has emerged as an important opportunity for improving clinical decision support across diverse healthcare environments.

Despite substantial advances in machine learning for critical care, generalization remains one of the principal barriers to translating artificial intelligence into routine clinical practice. Models developed and validated using data from a single institution or patient population frequently experience performance degradation when applied to different hospitals, healthcare systems, or demographic groups because of variations in patient characteristics, clinical workflows, data quality, and acquisition protocols [3,5–9]. Consequently, many AI systems that demonstrate excellent performance during retrospective evaluation fail to achieve comparable reliability in external validation, limiting clinician trust and widespread adoption [6–9]. Addressing these challenges requires learning physiological representations that remain robust across heterogeneous clinical environments rather than optimizing models solely for performance within individual datasets.

An often-overlooked challenge underlying this lack of generalizability is temporal resolution mismatch. Physiological data are collected at widely varying sampling frequencies across intensive care units because of differences in bedside monitoring infrastructure, clinical workflows, staffing, and resource availability [3,4,19]. As a result, the same physiological variable may be available as high-frequency continuous monitoring in one institution but only as intermittently recorded observations in another. Despite this variability, existing physiological representation learning and foundation model approaches are typically pretrained and evaluated at a fixed temporal resolution, implicitly assuming that the monitoring frequency encountered during deployment matches that used during model development [5,11–13]. Consequently, when models are applied to healthcare environments with different monitoring frequencies, they often require retraining or recalibration, increasing computational cost and limiting their scalability and clinical utility [3,9]. Whether a single pretrained physiological representation can remain robust across heterogeneous temporal resolutions without additional representation learning remains largely unexplored.

Self-supervised representation learning has emerged as a powerful paradigm for learning transferable representations from large collections of unlabeled biomedical data. Inspired by advances in natural language processing and foundation models, recent approaches have demonstrated that pretraining on large-scale clinical and physiological datasets can substantially improve performance across multiple downstream prediction tasks while reducing reliance on manual annotation [5,11–16]. Transformer-based architectures, including BEHRT for longitudinal electronic health records and large clinical language models such as GatorTron, have further demonstrated the potential of pretrained representations to support diverse clinical applications [12,13]. Similarly, self-supervised learning methods for physiological time series have focused on learning generalized representations from continuous monitoring data that can be transferred across downstream prediction tasks [11]. However, despite these advances, representation transfer has been evaluated almost exclusively across clinical tasks or datasets, while the ability of learned physiological representations to remain robust across heterogeneous temporal resolutions has received little attention. This represents an important gap because deployment environments frequently differ in monitoring frequency from those used during model development.

Progress in physiological representation learning has been driven largely by the availability of large, openly accessible critical care databases such as MIMIC-III, eICU, and the Pediatric Intensive Care (PIC) database, which have become essential resources for developing and benchmarking clinical AI models [10,17,18]. However, publicly available pediatric intensive care datasets containing longitudinal high-frequency physiological monitoring remain scarce, particularly in low- and middle-income countries where disease burden, resource constraints, and monitoring practices differ substantially from those in high-income healthcare systems [4,19,20]. The limited availability of such datasets has constrained the development, reproducibility, and external validation of pediatric physiological representation learning methods while restricting opportunities to investigate model robustness across diverse clinical environments. To address this gap, we developed SafeICU, an open-access pediatric intensive care database comprising longitudinal high-resolution physiological signals together with laboratory measurements, treatment records, microbiology, and clinical outcomes. By releasing both the dataset and pretrained PhysioStack representation models, we aim to provide a community resource that supports reproducible research, external benchmarking, and the development of generalizable physiological foundation models for pediatric critical care.

In this study, we present PhysioStack, a self-supervised physiological representation learning framework pretrained on SafeICU, a large-scale pediatric intensive care database comprising high-resolution longitudinal physiological signals and multimodal clinical data. We investigate whether physiological representations learned from high-frequency monitoring can be transferred across clinically relevant temporal resolutions without additional pretraining and evaluate their robustness across independent pediatric and adult intensive care cohorts. We further demonstrate the utility of these pretrained representations for downstream clinical prediction through early shock prediction and publicly release both the SafeICU database and pretrained PhysioStack models to facilitate reproducible research, external benchmarking, and future advances in physiological representation learning for critical care.

Our work makes four principal contributions. First, we introduce PhysioStack, a self-supervised physiological representation learning framework pretrained on high-resolution longitudinal physiological signals from the SafeICU database. Second, we demonstrate that pretrained physiological representations can be transferred across heterogeneous temporal resolutions without additional pretraining, addressing a previously unexplored challenge in physiological representation learning. Third, we show that these representations generalize across independent pediatric and adult intensive care cohorts and support accurate downstream clinical prediction through early shock prediction. Finally, we publicly release both the SafeICU database and pretrained PhysioStack models to enable reproducible research, external benchmarking, and future development of transferable physiological representation learning methods for critical care.

## 2. Methodology

### 2.1 Study design and data source

This study was conducted as a retrospective observational cohort study using data from SafeICU, a pediatric intensive care unit (PICU) database curated from routinely collected clinical data at AIIMS, New Delhi, India. The analyses reported here used data collected between January 2016 and December 2024; data collection for the SafeICU database is ongoing, with future updates planned.

SafeICU was developed to address the limited availability of high-resolution pediatric intensive care datasets suitable for self-supervised representation learning and the development of generalizable artificial intelligence models. Compared with adult intensive care populations, pediatric patients exhibit age-dependent physiological ranges, rapidly evolving developmental physiology, and heterogeneous disease trajectories, making direct transfer of adult-trained models challenging. Although publicly available resources such as MIMIC-III and eICU have substantially advanced critical care research, they primarily represent adult populations, while pediatric databases containing continuous high-frequency physiological monitoring remain scarce. By providing longitudinal physiological monitoring collected during routine clinical care in a tertiary-care hospital in India, SafeICU complements existing critical care databases and enables systematic investigation of physiological representation learning, temporal robustness, cross-cohort generalization, and future multimodal clinical AI applications in pediatric critical care.

The database integrates longitudinal patient-level demographic information, diagnoses, treatments, laboratory measurements, microbiology results, clinical notes, outcomes, and continuously recorded physiological signals. Continuous vital signs, including heart rate (HR), respiratory rate (RR), oxygen saturation (SpO_2_), arterial blood pressure (ABP), and temperature, were acquired directly from bedside monitoring systems at 15-second temporal resolution. This combination of high-frequency physiological monitoring and comprehensive clinical information provides a unique resource for developing and evaluating self-supervised representation learning methods while supporting future multimodal clinical AI research. Each record in SafeICU corresponds to a single PICU admission.

The study protocol was reviewed and approved by the Ethics Committee of AIIMS, New Delhi (approval numbers IEC/NP-211/08·05·2015 and IEC-787/07.10.2022, RP-14/2022). The requirement for individual informed consent was waived because the study involved retrospective analysis of routinely collected clinical data, and all data were anonymized before use. This observational study was conducted and reported in accordance with the STROBE (Strengthening the Reporting of Observational Studies in Epidemiology) guidelines [21]. As the data used were routinely collected during standard clinical care, reporting was also guided by the RECORD (REporting of studies Conducted using Observational Routinely-collected health Data) statement [22].

To evaluate the generalizability of the learned physiological representations beyond the pediatric cohort and institutional setting, adult intensive care data were obtained from the Medical Information Mart for Intensive Care III (MIMIC-III) database [18] and the eICU Collaborative Research Database [10], both publicly available through PhysioNet. These datasets were used exclusively for external evaluation of representation transferability and were not included in model pretraining.

### 2.2 Data elements and variables

#### Data integration across clinical streams

SafeICU integrates routinely collected structured clinical data and high-frequency physiological time-series data into a unified relational database. The database schema was designed to be broadly compatible with the MIMIC-IV framework to facilitate usability and comparison with established critical care datasets. Data were organized into core tables, including patient demographics (PATIENT), admissions (ADMISSION), treatment chart documentation (NOTE EVENTS), continuously recorded physiological signals (VITAL SIGNS), laboratory results (LAB EVENTS), and microbiology results (MICROBIOLOGY EVENTS). Records across tables were linked using a unique patient identifier (SUBJECT_ID) and an admission-level identifier. The integrated database was designed to support both conventional clinical research and modern machine-learning applications by preserving longitudinal patient trajectories across multiple clinical modalities. The availability of synchronized physiological monitoring, laboratory measurements, treatment documentation, microbiology results, and clinical outcomes enables investigations ranging from predictive modelling to self-supervised representation learning and future multimodal foundation models.

Following extraction from source hospital systems, records were harmonized and filtered to ensure internal consistency across clinical streams. Basic data quality checks were performed, including validation of age ranges, removal of physiologically unacceptable values, and handling of extreme outliers. Because time ranges differed across source systems, integration was performed using SUBJECT_ID-based linkage to retain clinically coherent patient episodes. An overview of the SafeICU database structure and integration workflow is provided in Figure 1, database characteristics are summarised in Supplementary Material 1, and a comparative summary of SafeICU against widely used publicly available critical care databases is provided in Supplementary Material 2.

**Figure 1.**
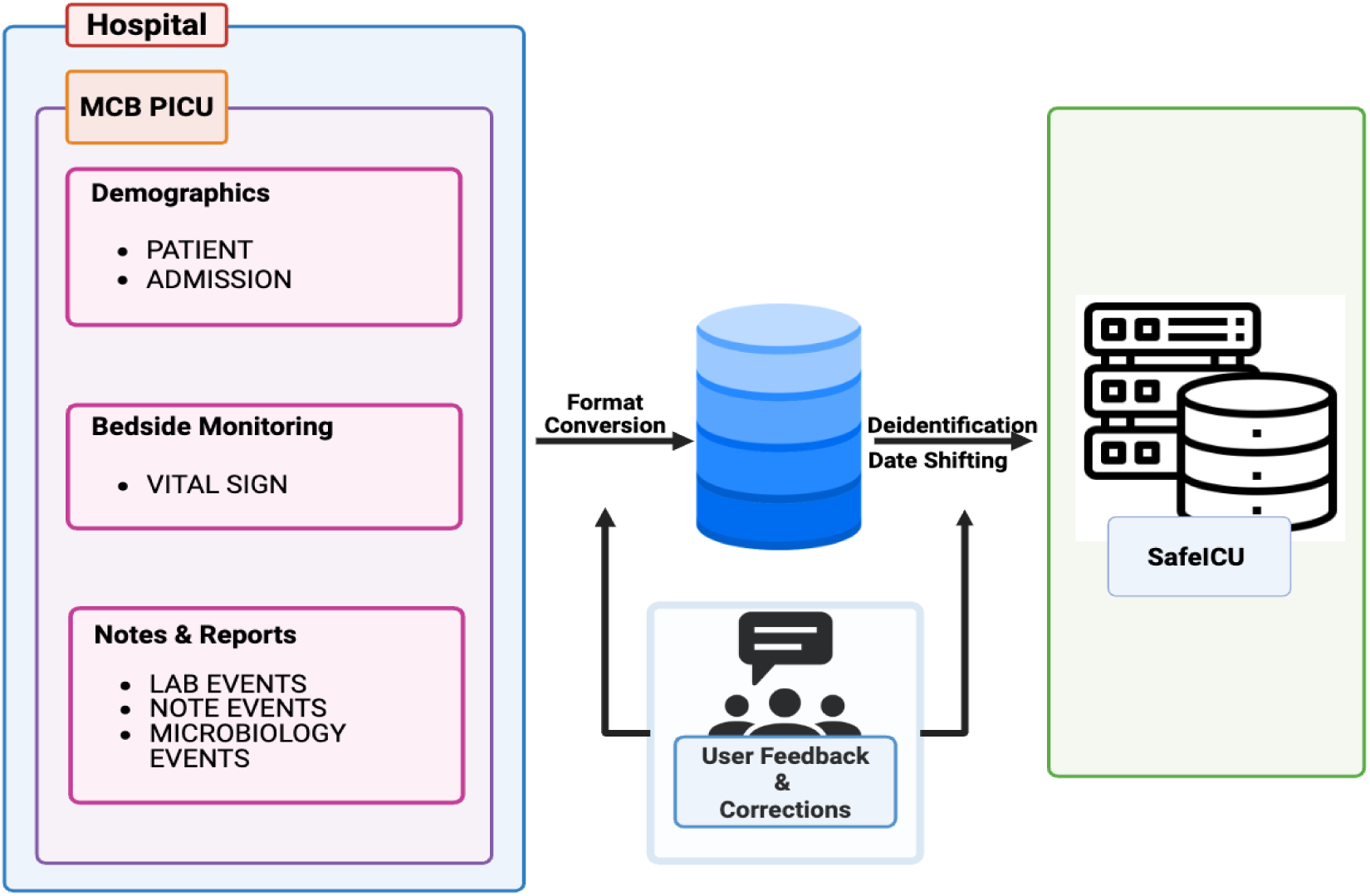
SafeICU database structure and integration workflow. Routinely collected clinical data streams were harmonised using unique patient identifiers to construct a unified de-identified research database.

#### Data anonymization

Before incorporation into the SafeICU database, all data were de-identified in accordance with Health Insurance Portability and Accountability Act (HIPAA) guidelines [23]. Structured de-identification was performed through the removal of direct identifiers, including patient names, clinician names, and other free-text identifiers. Dates were consistently shifted forward for each patient using a randomly generated offset, preserving within-patient temporal intervals while preventing identification through calendar dates.

Unique identifiers and institutional codes were pseudonymised using a secure hashing approach (PBKDF2 with HMAC-SHA256), ensuring that original identifiers could not be recovered from the released dataset. De-identification procedures were applied uniformly across all clinical streams before database integration. Because the cohort is restricted to patients younger than 18 years, additional age-hiding procedures used in adult datasets (eg, masking ages >89 years) were not required.

### 2.3 Study population

The source population comprised all pediatric patients aged below 18 years who were admitted to the PICU during the study period. The analytic cohort used for vitals-based representation learning included PICU admissions with available continuous physiological monitoring data for HR, ABP, RR, and SpO_2_.

Admissions were excluded if any of the required vital signs were entirely missing or if the duration of available physiological data was insufficient for analysis, defined as fewer than six recorded data points for any vital sign. These criteria were applied to ensure that included admissions had adequate longitudinal data to support temporal aggregation across monitoring intervals. Although the SafeICU database includes additional clinical data streams, such as laboratory, microbiology, and treatment data, inclusion in the cohort for this study was based solely on the availability of continuous vital-sign data.

For patients with multiple PICU admissions, each stay was included independently. In addition, interruptions in physiological monitoring exceeding 24 hours were considered as a new ICU stay and were treated as separate admissions.

For downstream evaluation, the pediatric cohort definition and outcome labeling were held consistent with a previously published study derived from the SafeICU database. Adult ICU cohorts used for external generalization analyses were derived from the MIMIC-III and eICU Collaborative Research Database and were restricted to patients with available continuous recordings of the same physiological variables.

### 2.4 Temporal resolution and aggregation strategy

Physiological signals in SafeICU were originally recorded at 15-second temporal resolution. Missingness in the continuous physiological time-series was assessed across the four primary vital signs (HR, RR, SpO_2_, and ABP). Admissions with extensive missingness were excluded; specifically, admissions were removed if overall missingness across the four vital sign streams exceeded 50%. Critical patient data for eligible patients in the development cohort were imputed using the Kalman filter. Imputation was performed independently for each physiological signal to preserve within-signal temporal structure.

To evaluate robustness under heterogeneous monitoring frequencies, we first segmented the continuous 15-second physiological time series into overlapping sliding windows, with a 30-minute overlap between consecutive segments to preserve temporal continuity and contextual information. Following this sharding step, coarser time-series representations were generated by aggregating the native 15-second signals into clinically relevant temporal intervals of 5, 10, 15, 30, and 60 minutes. For each vital sign, values within each aggregation window were summarised using the median to reduce sensitivity to outliers and transient artefacts. Windows with insufficient recorded values were treated as missing.

To simulate real-world deployment scenarios where monitoring is intermittently recorded at a lower frequency, models trained at one temporal resolution were evaluated on data aggregated at different resolutions without retraining. This enabled systematic assessment of temporal-resolution mismatch. Temporal aggregation was performed independently for each admission after segmentation of continuous monitoring into distinct PICU stays.

### 2.5 Symbolic encoding and self-supervised representation learning

To enable self-supervised learning on physiological time-series data, continuous vital sign measurements were transformed into discrete symbolic sequences. For each physiological variable (HR, RR, SpO_2_, and ABP), values were discretised into a fixed number of bins to generate integer-valued tokens. Discretisation was performed using a clustering-based approach, where cluster centroids defined data-driven physiological state boundaries.

Self-supervised representation learning was performed using a BERT-base uncased transformer architecture trained from scratch with an MLM objective [14]. During training, 15% of tokens within each physiological sequence were randomly masked, and the model was trained to predict the masked tokens using surrounding contextual information. Sequences were constructed from temporally ordered symbolic tokens derived from aggregated vital sign time series, enabling the model to learn contextual dependencies and temporal patterns across physiological trajectories.

The model was optimised using a cross-entropy loss between predicted and true masked tokens. After pretraining, the learned embeddings were extracted as fixed-dimensional representations for downstream evaluation, including temporal resolution transfer experiments and early shock prediction. Hyperparameters for pretraining were optimised using Optuna through Bayesian optimisation to minimise validation loss (Supplementary Material 3). The final model configuration was selected based on the best-performing trial on the validation set.

### 2.6 Downstream evaluation: early shock prediction

To evaluate whether the learned physiological representations retained clinically meaningful discriminative information under temporal resolution transfer, we performed a downstream early shock prediction task. Shock prediction was selected as an illustrative application because it is clinically relevant in intensive care and has been widely used in prior machine learning studies for evaluating the clinical utility of learned physiological representations. In this study, shock prediction was used to assess the stability of pretrained embeddings across temporal resolutions and patient cohorts rather than to characterise the full clinical determinants of shock.

Shock onset was defined using the Shock Index (SI), calculated as the ratio of heart rate to systolic blood pressure, which has been validated for bedside risk stratification and prognostication [24,25]. In adult populations, SI values greater than 0.7 are commonly used to indicate shock. Given that SafeICU is a pediatric data resource, age-adjusted SI thresholds were applied to account for physiological variation across developmental stages. Shock was defined as SI >2.3 (≤3 months), >1.7 (4-6 months), >1.5 (7-12 months), >1.2 (13-36 months), >1.15 (37-72 months), >0.95 (73-144 months), and >0.77 (>144 months). The prediction horizon was set to 8 hours before shock onset [26]. The pediatric cohort definition, including inclusion and exclusion criteria, data splits, and outcome labeling procedure, is described in Supplementary Material 4 and was held constant across all analyses to enable direct comparison while isolating the effect of temporal resolution.

Pretrained physiological embeddings were extracted independently for each vital sign using the corresponding pretrained representation model. For temporal resolution transfer experiments, embeddings were generated using a model pretrained at 5-minute resolution and applied to data aggregated at coarser resolutions (10-60 minutes) without retraining. Embeddings from all physiological signals were concatenated to form a unified feature vector. Dimensionality reduction was performed using principal component analysis, and age and sex were included as additional covariates. No fine-tuning of the pretrained representation models was performed during downstream classifier training.

Several standard classifiers were evaluated, including logistic regression, support vector machines, random forests, and gradient boosting. Model selection was performed using five-fold cross-validation on the training set. Based on cross-validated performance, gradient boosting was selected as the final classifier. The selected model was retrained on the combined training and validation sets and evaluated on a held-out test set. The same feature construction and evaluation pipeline was applied to both pediatric and adult ICU cohorts to enable direct comparison of downstream performance under temporal resolution mismatch and across patient populations.

### 2.7 Evaluation metrics and statistical analysis

Model performance was summarised descriptively across physiological signals, temporal resolutions, and patient cohorts. For self-supervised representation learning, masked modelling performance was evaluated using MLM accuracy and validation loss for each vital sign and aggregation interval. For downstream early shock prediction, discrimination was assessed using the area under the receiver operating characteristic curve (AUROC) and area under the precision-recall curve (AUPRC), with performance compared across temporal resolutions to quantify the effect of temporal-resolution mismatch at inference. All analyses were performed using Python-based scientific computing libraries, including PyTorch for model training and scikit-learn for downstream classification and evaluation.

## 3 Results

### 3.1 SafeICU cohort characteristics

The pretraining cohort comprised 621 unique pediatric patients from the SafeICU database, corresponding to 984 PICU stays after temporal segmentation of physiological monitoring streams, where interruptions in recording exceeding 24 hours were treated as separate stays. This analytic cohort represents the subset of SafeICU admissions with sufficient continuous physiological monitoring data across the four primary vital signs (HR, RR, SpO_2_, and ABP) after missingness-based filtering and imputation. Data were partitioned at the patient level into training (434 patients), validation (63 patients), and test (124 patients) sets for language model training, hyperparameter optimisation, and masked modelling evaluation, respectively.

### 3.2 Validation of self-supervised pretraining

Before evaluating representation transferability, we first verified that the self-supervised MLM successfully learned meaningful physiological representations. Hyperparameter optimization substantially improved training stability, producing faster convergence and lower validation loss than the baseline configuration (Figure 2).

**Figure 2.**
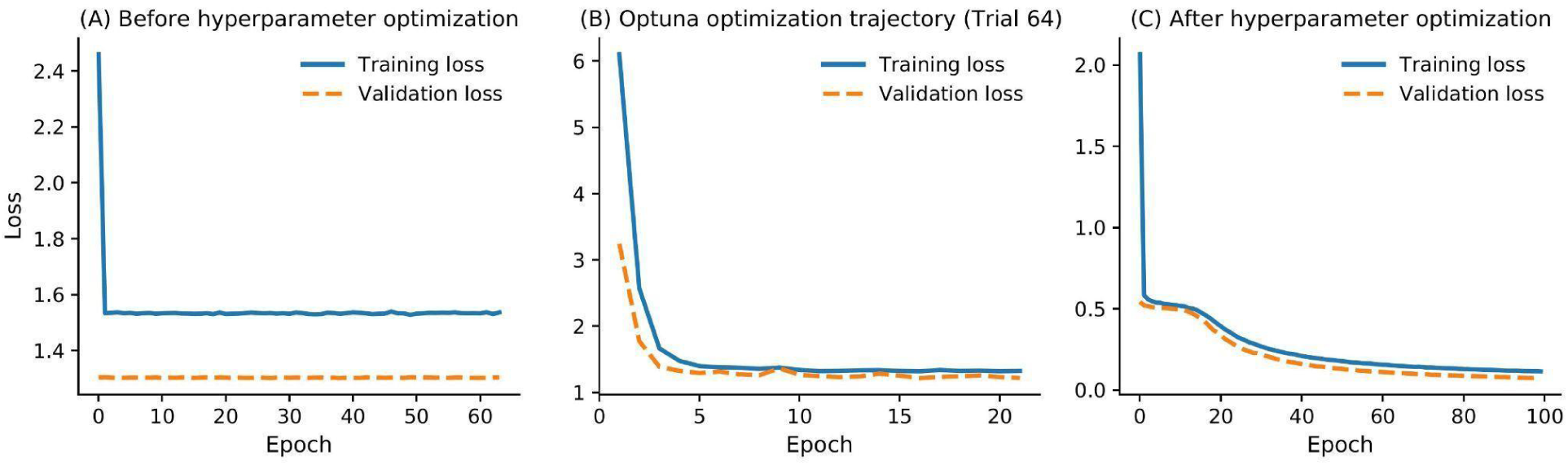
Training and validation loss during masked modelling pretraining. Loss trajectories for MLM pretraining on heart rate sequences at 5-minute resolution. (A) Baseline hyperparameters. (B) Best-performing Optuna trial (Trial 64). (C) Final model trained using optimized hyperparameters, demonstrating faster convergence and lower validation loss.

Optimized models consistently achieved higher MLM accuracy across all physiological variables than baseline models (Table 1). At the native 5-minute resolution, median MLM accuracy increased from 0.42 to 0.79 for HR, 0.45 to 0.74 for RR, 0.55 to 0.84 for SpO_2_, and 0.50 to 0.92 for ABP. As expected, MLM accuracy gradually decreased with increasing temporal aggregation because coarser sampling contains less temporal information. Similar trends were observed in the external adult cohort (Supplementary Material 5).

**Table 1.** MLM accuracy across temporal resolutions in the SafeICU pediatric cohort. Median (IQR) MLM accuracy for HR, RR, SpO_2_, and ABP at aggregation intervals from 5 to 60 minutes. Baseline and Optuna-optimized results are shown for 5-minute training; all other resolutions reflect optimized models.

| Temporal resolution | Training configuration | MLM accuracy, HR (Median [IQR]) | MLM accuracy, RR (Median [IQR]) | MLM accuracy, SpO<sub>2</sub> (Median [IQR]) | MLM accuracy, ABP (Median [IQR]) |
| --- | --- | --- | --- | --- | --- |
| 5 minutes | Baseline hyperparameters | 0.42 (0.29-0.53) | 0.45 (0.34-0.58) | 0.55 (0.42-0.68) | 0.5 (0.39-0.66) |
| 5 minutes | Optimized hyperparameters | 0.79 (0.68-0.87) | 0.74 (0.63-0.84) | 0.84 (0.76-0.89) | 0.92 (0.74-0.97) |
| 10 minutes | Optimized hyperparameters | 0.74 (0.63-0.84) | 0.67 (0.55-0.79) | 0.8 (0.71-0.87) | 0.63 (0.51-0.76) |
| 15 minutes | Optimized hyperparameters | 0.68 (0.56-0.79) | 0.66 (0.55-0.76) | 0.76 (0.63-0.87) | 0.84 (0.71-0.95) |
| 30 minutes | Optimized hyperparameters | 0.61 (0.42-0.76) | 0.37 (0.24-0.5) | 0.42 (0.32-0.54) | 0.8 (0.67-0.92) |
| 60 minutes | Optimized hyperparameters | 0.53 (0.39-0.67) | 0.55 (0.42-0.71) | 0.4 (0.29-0.5) | 0.34 (0.21-0.48) |

Abbreviations: ABP, arterial blood pressure; HR, heart rate; IQR, interquartile range; MLM, masked language modeling; RR, respiratory rate; SpO<sub>2</sub>, oxygen saturation.

These experiments establish that PhysioStack successfully learned stable physiological representations suitable for downstream evaluation. The following analyses therefore focus on the central question of this study, whether representations learned from high-frequency monitoring remain transferable across temporal resolutions and patient populations without retraining.

### 3.3 Temporal resolution transfer and cross-cohort generalization

To evaluate the robustness of the learned physiological representations, we investigated whether a single PhysioStack model pretrained at the highest temporal resolution (5 minutes) could be transferred to coarser temporal resolutions without additional retraining, and whether these representations generalized across independent ICU cohorts. Figure 3 summarizes the results for both pediatric and adult cohorts.

**Figure 3.**
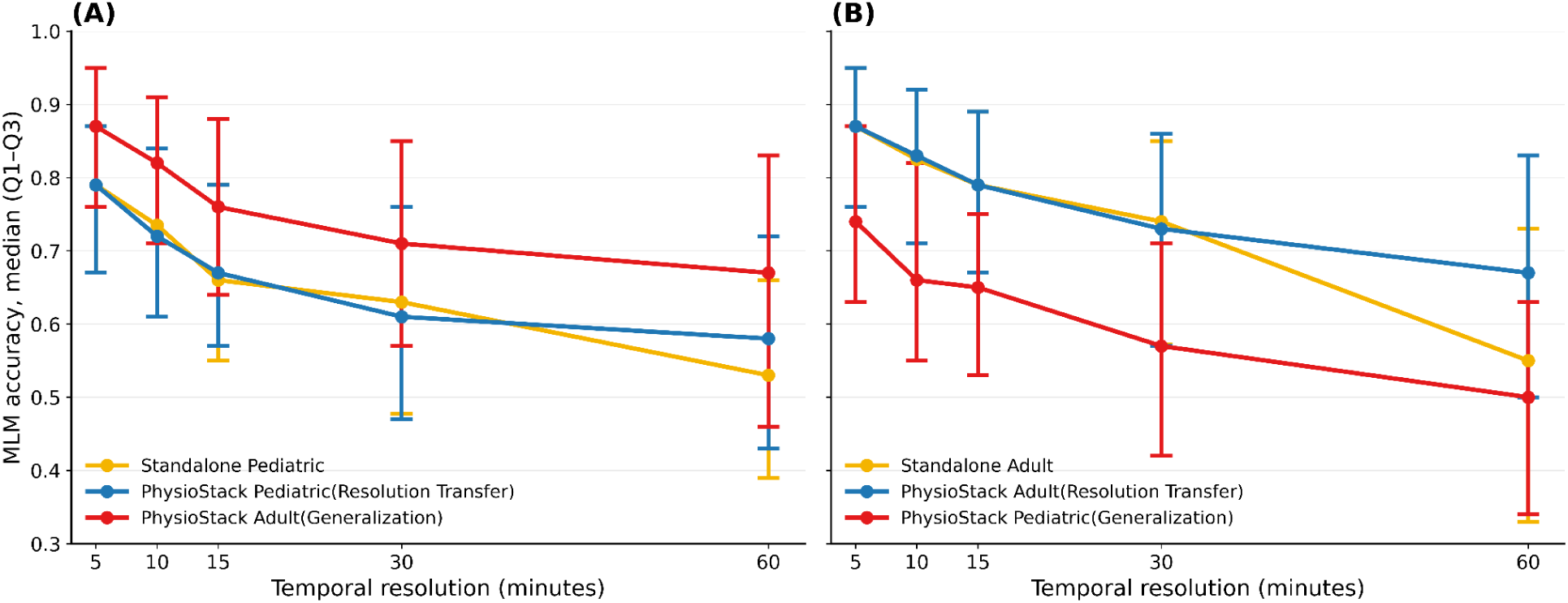
Temporal resolution transfer and cross-cohort generalization of PhysioStack representations. (A) The yellow curve represents independently trained resolution-specific pediatric baseline models, the blue curve represents the 5-minute PhysioStack pediatric model evaluated across pediatric temporal resolutions, and the red curve represents the same pretrained pediatric model evaluated on the adult cohort. (B) The yellow curve represents independently trained resolution-specific adult baseline models, the blue curve represents the 5-minute PhysioStack adult model evaluated across adult temporal resolutions, and the red curve represents the same pretrained adult model evaluated on the pediatric cohort. Points indicate the median MLM accuracy, while error bars denote the interquartile range (Q1-Q3).

As shown in Figure 3A, the PhysioStack model pretrained on 5-minute pediatric physiological signals consistently preserved high MLM accuracy across all temporal resolutions without requiring additional retraining. The transferred model achieved median accuracies of 0.79, 0.72, 0.67, 0.61, and 0.58 when evaluated on 5-, 10-, 15-, 30-, and 60-minute pediatric data, respectively. Across all resolutions, its performance remained comparable to that of independently trained resolution-specific pediatric models. These findings demonstrate that representations learned from high-resolution physiological data can be effectively transferred across substantially coarser monitoring intervals without retraining, highlighting the temporal robustness of the learned representations. Furthermore, when the same pretrained pediatric model was directly evaluated on the independent adult eICU cohort, it achieved median MLM accuracies of 0.87, 0.82, 0.76, 0.71, and 0.67 across corresponding temporal resolutions, demonstrating strong cross-cohort generalization despite differences in patient populations and clinical environments.

A complementary experiment is presented in Figure 3B, where models were pretrained using the adult eICU cohort. Similar trends were observed. The independently trained adult baseline achieved median MLM accuracies of 0.87, 0.83, 0.79, 0.74, and 0.55 across increasing temporal resolutions. The single 5-minute PhysioStack adult model preserved comparable performance after temporal aggregation, yielding median accuracies of 0.87, 0.83, 0.79, 0.73, and 0.67. When evaluated on the independent SafeICU pediatric cohort without additional optimization, the pretrained adult model achieved median accuracies of 0.74, 0.66, 0.65, 0.57, and 0.50, confirming bidirectional transferability of the learned representations across heterogeneous ICU populations.

Together, these findings demonstrate that PhysioStack learns temporal representations that are robust to substantial reductions in sampling frequency while also generalizing effectively across independent pediatric and adult ICU cohorts. Rather than requiring separate pretraining for each temporal resolution or clinical population, a single high-resolution pretrained model can be transferred across multiple temporal granularities and patient cohorts with minimal degradation in representation quality.

### 3.4 Downstream evaluation: Early warning system for predicting shock

To evaluate whether the learned physiological representations retained clinically meaningful information beyond the pretraining task, we assessed their utility for early shock prediction using the SafeICU pediatric cohort. Under five-fold cross-validation, the model demonstrated strong discrimination, with a median AUROC of 0.87 (IQR 0.86-0.88) and AUPRC of 0.92 (0.91-0.93) (Supplementary Material 6). When evaluated under temporal resolution mismatch without retraining, predictive performance remained stable across coarser aggregation intervals from 10 to 60 minutes, with AUROC ranging from 0.82 to 0.86 and AUPRC from 0.91 to 0.94, indicating that representations learned from high-resolution physiological monitoring remained informative despite substantial temporal downsampling.

Across all evaluated temporal resolutions, classification performance remained consistently high (Table 2). Accuracy ranged from 0.79 to 0.84, while F1 scores remained between 0.85 and 0.90. Sensitivity was consistently high (0.93-0.99) across all resolutions, demonstrating reliable identification of patients who subsequently developed shock. AUROC remained stable between 0.81 (95% CI, 0.77-0.85) and 0.86 (95% CI, 0.82-0.89), with the highest discrimination observed at 15-minute resolution. Similarly, AUPRC varied only modestly across temporal resolutions, ranging from 0.91 (95% CI, 0.88-0.94) to 0.94 (95% CI, 0.91-0.96). Model calibration also remained consistent, with Brier scores between 0.13 and 0.17, indicating good agreement between predicted probabilities and observed outcomes.

**Table 2.** Downstream early shock prediction performance across temporal resolutions using PhysioStack representations. Performance of the gradient boosting classifier trained using embeddings extracted from the pretrained 5-minute PhysioStack model and evaluated on pediatric physiological data aggregated to 5-, 10-, 15-, 30-, and 60-minute temporal resolutions. Performance metrics are summarized from bootstrap resampling. Values are reported as mean ± standard deviation, except AUROC and AUPRC, which are reported with 95% confidence intervals.

| Metric | 5 minutes | 10 minutes | 15 minutes | 30 minutes | 60 minutes |
| --- | --- | --- | --- | --- | --- |
| Accuracy | 0.83 ± 0.017 | 0.83 ± 0.017 | 0.82 ± 0.018 | 0.79 ± 0.019 | 0.84 ± 0.016 |
| Sensitivity (Recall) | 0.95 ± 0.011 | 0.95 ± 0.011 | 0.94 ± 0.013 | 0.93 ± 0.013 | 0.99 ± 0.005 |
| Specificity | 0.47 ± 0.044 | 0.47 ± 0.044 | 0.48 ± 0.043 | 0.27 ± 0.039 | 0.43 ± 0.043 |
| F1 score | 0.89 ± 0.012 | 0.89 ± 0.012 | 0.88 ± 0.012 | 0.85 ± 0.013 | 0.90 ± 0.011 |
| MCC | 0.51 ± 0.044 | 0.51 ± 0.045 | 0.48 ± 0.047 | 0.27 ± 0.050 | 0.57 ± 0.038 |
| AUROC (95% CI) | 0.82(0.77-0.86) | 0.82(0.77-0.86) | 0.86(0.82-0.89) | 0.81(0.77-0.85) | 0.82(0.78-0.86) |
| AUPRC (95% CI) | 0.91(0.88-0.94) | 0.91(0.87-0.94) | 0.94(0.91-0.96) | 0.92(0.90-0.95) | 0.91(0.88-0.94) |
| Brier Score | 0.14 ± 0.013 | 0.13 ± 0.012 | 0.13 ± 0.012 | 0.17 ± 0.012 | 0.14 ± 0.013 |

Abbreviations: MCC, Matthews Correlation Coefficient; AUROC, area under the receiver operating characteristic curve; AUPRC, area under the precision-recall curve.

These observations are further illustrated in Figure 4. Receiver operating characteristic curves (Figure 4A) showed comparable discrimination across all temporal resolutions, with closely overlapping ROC curves and consistently high AUROC values. Precision-recall curves (Figure 4B) likewise demonstrated stable performance despite temporal aggregation, with AUPRC remaining above 0.90 for every evaluated resolution. Calibration plots (Figure 4C) showed that predicted probabilities closely followed the ideal calibration line, indicating that probability estimates remained well calibrated after transferring representations from the pretrained 5-minute model to coarser monitoring frequencies.

**Figure 4.**
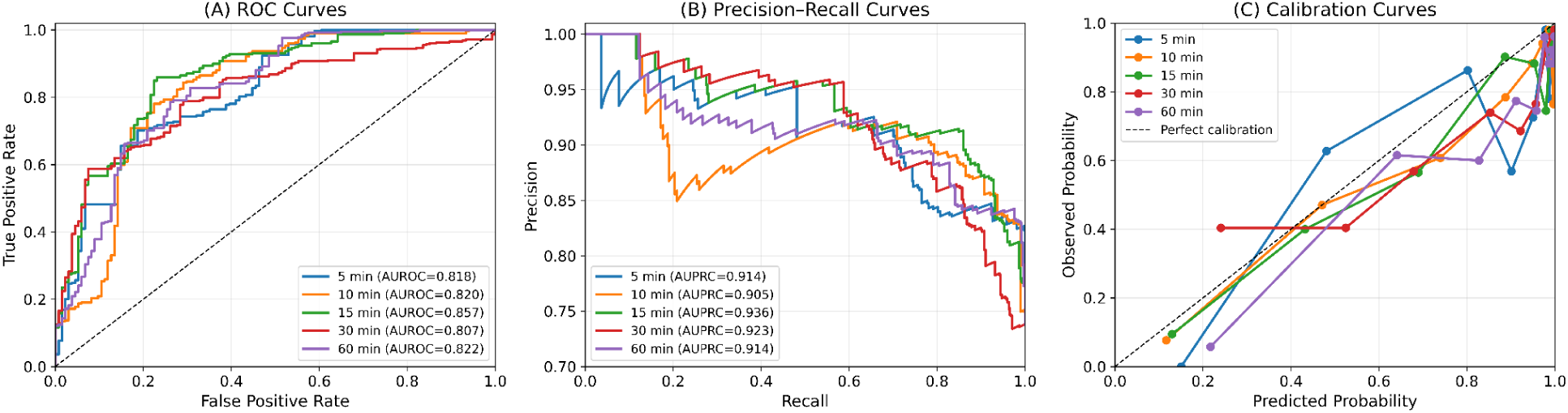
Downstream performance of PhysioStack representations for early shock prediction across temporal resolutions. Gradient boosting classifiers were trained using embeddings extracted from the pretrained 5-minute PhysioStack model and evaluated on pediatric physiological data aggregated to 5-, 10-, 15-, 30-, and 60-minute temporal resolutions. (A) Receiver operating characteristic curves comparing the trade-off between sensitivity and false positive rate for shock prediction across temporal resolutions. (B) Precision-Recall curves illustrate the balance between precision and recall, providing a more informative assessment for the imbalanced shock prediction task. (C) Calibration curves comparing predicted probabilities with observed event frequencies to assess the reliability of model probability estimates across temporal resolutions.

### 3.5 Cross-Cohort Generalization: Early Warning System for Predicting Shock

To assess whether the learned physiological representations generalized beyond the training cohort, we evaluated downstream shock prediction on an independent ICU cohort using embeddings extracted from the pretrained PhysioStack pediatric model without modifying the representation model. Performance was assessed across temporal resolutions ranging from 5 to 60 minutes (Table 3).

**Table 3.** Cross-cohort downstream early shock prediction performance using transferred PhysioStack representations. Performance of the gradient boosting classifier trained using embeddings extracted from the pretrained 5-minute pediatric PhysioStack model and evaluated on the independent adult eICU cohort across 5-, 10-, 15-, 30-, and 60-minute temporal resolutions. Results are summarized from bootstrap resampling and reported as mean ± standard deviation, except AUROC and AUPRC, which are reported with 95% confidence intervals.

| Metric | 5 minutes | 10 minutes | 15 minutes | 30 minutes | 60 minutes |
| --- | --- | --- | --- | --- | --- |
| Accuracy | 0.85 ± 0.003 | 0.89 ± 0.003 | 0.89 ± 0.002 | 0.90 ± 0.002 | 0.91 ± 0.002 |
| Sensitivity (Recall) | 0.92 ± 0.003 | 0.93 ± 0.002 | 0.93 ± 0.002 | 0.93 ± 0.002 | 0.93 ± 0.002 |
| Specificity | $0.71 \pm 0.006$ | $0.80 \pm 0.006$ | $0.81 \pm 0.005$ | $0.84 \pm 0.005$ | $0.86 \pm 0.005$ |
| F1 score | $0.89 \pm 0.002$ | $0.92 \pm 0.002$ | $0.92 \pm 0.002$ | $0.93 \pm 0.002$ | $0.93 \pm 0.002$ |
| MCC | $0.65 \pm 0.006$ | $0.73 \pm 0.006$ | $0.74 \pm 0.006$ | $0.76 \pm 0.005$ | $0.78 \pm 0.005$ |
| AUROC (95% CI) | 0.91(0.90-0.91) | 0.93(0.92-0.93) | 0.93(0.92-0.93) | 0.94(0.94-0.95) | 0.95(0.95-0.95) |
| AUPRC (95% CI) | 0.94(0.94-0.95) | 0.95(0.95-0.96) | 0.95(0.95-0.96) | 0.96(0.95-0.96) | 0.97(0.96-0.97) |
| Brier Score | $0.13 \pm 0.001$ | $0.11 \pm 0.001$ | $0.11 \pm 0.001$ | $0.09 \pm 0.001$ | $0.09 \pm 0.001$ |

Abbreviations: MCC, Matthews Correlation Coefficient; AUROC, area under the receiver operating characteristic curve; AUPRC, area under the precision-recall curve.

The pretrained representations demonstrated excellent cross-cohort transferability, achieving consistently high predictive performance across all temporal resolutions. Accuracy improved progressively from 0.85 ± 0.003 at 5-minute resolution to 0.91 ± 0.002 at 60-minute resolution, while sensitivity remained consistently high (0.92-0.93) across all evaluation settings, indicating reliable identification of patients who subsequently developed shock. Specificity likewise increased with temporal aggregation, rising from 0.71 ± 0.006 to 0.86 ± 0.005, accompanied by corresponding improvements in MCC from 0.65 ± 0.006 to 0.78 ± 0.005.

Discrimination performance remained consistently strong across all temporal resolutions. AUROC increased from 0.91 (95% CI, 0.90-0.91) at 5-minute resolution to 0.95 (95% CI, 0.95-0.95) at 60-minute resolution, while AUPRC improved from 0.94 (95% CI, 0.94-0.95) to 0.97 (95% CI, 0.96-0.97). Model calibration also remained stable, with Brier scores decreasing from 0.13 ± 0.001 to 0.09 ± 0.001, indicating increasingly accurate probability estimates across progressively coarser temporal resolutions.

These findings are further illustrated in Figure 5. Receiver operating characteristic curves (Figure 5A) showed excellent discrimination across all temporal resolutions, with AUROC values consistently exceeding 0.90. Precision-Recall curves (Figure 5B) similarly demonstrated robust predictive performance, maintaining AUPRC values above 0.94 across all evaluated resolutions. Calibration plots (Figure 5C) showed close agreement between predicted probabilities and observed outcomes, indicating well-calibrated risk estimates irrespective of temporal resolution.

**Figure 5.**
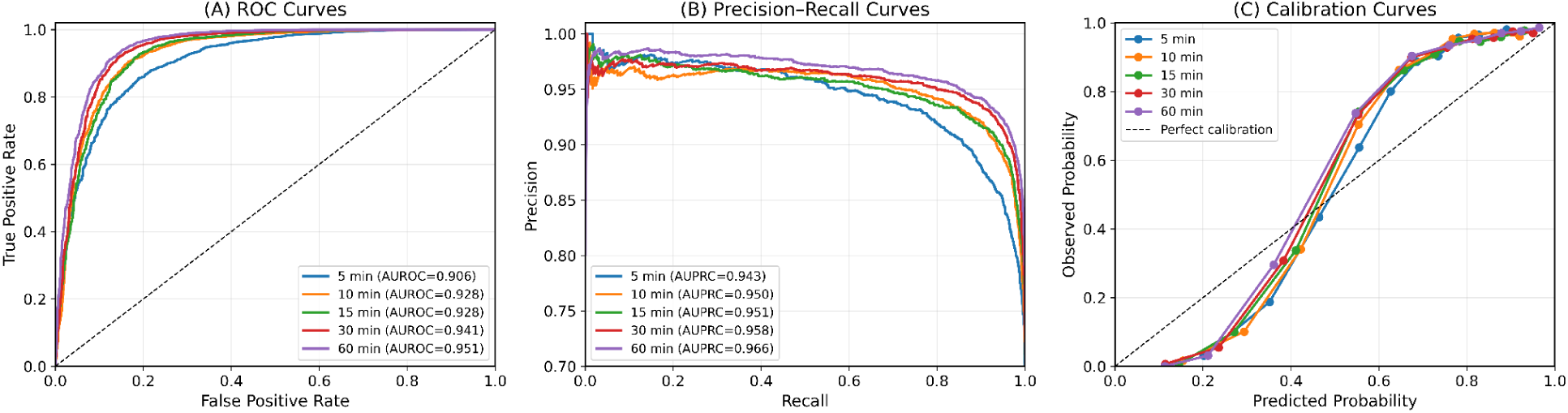
Cross-cohort downstream early shock prediction using transferred pediatric PhysioStack representations. Gradient boosting classifiers trained using embeddings extracted from the pretrained 5-minute pediatric PhysioStack model were evaluated on the independent adult eICU cohort across 5-, 10-, 15-, 30-, and 60-minute temporal resolutions. (A) Receiver operating characteristic (ROC) curves illustrate the discrimination performance of shock prediction models across temporal resolutions. (B) Precision-Recall (PR) curves showing the relationship between precision and recall for the imbalanced shock prediction task. (C) Calibration curves comparing predicted probabilities with observed shock outcomes to assess the reliability of probability estimates across temporal resolutions.

Collectively, these results demonstrate that PhysioStack representations learned from high-resolution physiological signals generalize effectively to an independent clinical cohort for downstream early shock prediction. Importantly, strong predictive performance was maintained across multiple temporal resolutions without retraining the representation model, highlighting the robustness and transferability of the learned physiological representations.

## Discussion

The present study makes three primary contributions toward advancing generalizable physiological AI for critical care. First, we demonstrate that self-supervised physiological representations learned from high-resolution monitoring can be effectively transferred across heterogeneous temporal resolutions while maintaining robust performance across independent pediatric and adult ICU cohorts without retraining. These findings establish temporal resolution transfer together with cross-cohort generalization as important evaluation paradigms for physiological representation learning [5,16,11]. Second, we demonstrate that the learned representations preserve clinically meaningful physiological information for downstream early shock prediction while maintaining robust performance under cross-cohort transfer, indicating that the learned representations generalize beyond the self-supervised pretraining objective. This highlights the potential of transferable physiological representations to support clinically relevant prediction across heterogeneous patient populations and healthcare settings [3,6,9]. Third, we publicly release the SafeICU database together with pretrained PhysioStack models, providing one of the most comprehensive openly available pediatric intensive care resources comprising longitudinal vital signs, laboratory measurements, treatment records, microbiology, and admission and discharge information. The scarcity of publicly available, high-resolution pediatric ICU datasets remains a major barrier to developing, validating, and externally benchmarking AI models for critically ill children, particularly in low-resource settings [4,17,19,20].

Recent advances in self-supervised learning and foundation models have demonstrated that large-scale pretraining enables the learning of transferable clinical representations that improve performance across multiple downstream prediction tasks [5,11–13,16]. Transformer-based models such as BEHRT extended representation learning to longitudinal electronic health records [12], while large-scale clinical foundation models have further highlighted the potential of pretrained representations for generalizable medical AI [5,13]. Similarly, recent self-supervised learning approaches for medical time series have primarily focused on improving downstream prediction by learning generalized representations from unlabeled physiological signals [11]. However, despite these advances, the transferability of learned representations has almost exclusively been evaluated across downstream tasks or external datasets, while implicitly assuming that the temporal resolution during deployment matches that used for model pretraining.

In contrast, monitoring frequency in real-world intensive care units is rarely standardized. Differences in bedside monitoring infrastructure, resource availability, staffing, and clinical workflows frequently result in heterogeneous sampling frequencies both within and across healthcare institutions [3,4,9,19]. Consequently, models trained using high-frequency physiological data often require retraining or redevelopment before deployment in settings with different monitoring practices. Our findings demonstrate that a single PhysioStack model pretrained on high-resolution physiological signals remains informative when applied to substantially coarser temporal resolutions without additional representation learning. Rather than treating temporal resolution mismatch as a limitation requiring separate models, our results suggest that temporal resolution transfer should be considered an essential dimension for evaluating physiological representation learning. This extends current evaluation paradigms from task transfer to temporal transfer, providing a more realistic assessment of model robustness for clinical deployment.

The next major finding of this study is the cross-cohort generalization achieved by the learned physiological representations. Limited external validity remains one of the principal barriers preventing the translation of clinical AI models into routine healthcare practice, as predictive performance often deteriorates when models are applied to institutions, patient populations, or acquisition protocols different from those used during development [6,8,9]. This challenge is particularly pronounced in pediatric critical care, where the scarcity of large, publicly available datasets frequently necessitates model development using single-center cohorts or transfer from adult populations [4,17,19]. PhysioStack maintained robust representation quality and downstream prediction performance when transferred between independent pediatric and adult ICU cohorts. Unlike conventional approaches, it achieved this without requiring cohort-specific retraining or recalibration. These findings suggest that the learned embeddings capture shared physiological dynamics that are preserved across age groups and healthcare systems, rather than encoding cohort-specific statistical characteristics. Such representation-level robustness complements the emerging vision of foundation models in medicine by demonstrating that generalizability should be evaluated not only across downstream tasks but also across independent clinical populations [5,16].

While temporal resolution transfer and cross-cohort generalization demonstrate the robustness of the learned representations, their clinical value ultimately depends on whether they preserve information relevant to downstream decision-making. To assess this, we evaluated the representations on early shock prediction, a clinically important task in pediatric critical care where timely intervention is essential for improving patient outcomes [1,2,24,27]. Unlike conventional approaches that typically train prediction models directly on task-specific features or require separate models for individual datasets and temporal resolutions [2,27], our downstream experiments used embeddings extracted from a single pretrained 5-minute PhysioStack model across all temporal resolutions. Despite substantial temporal aggregation and cross-cohort transfer, predictive performance remained consistently robust, indicating that the learned representations retained clinically meaningful physiological information beyond the self-supervised pretraining objective. These findings suggest that PhysioStack captures fundamental physiological dynamics associated with early clinical deterioration rather than learning representations optimized only for the masked language modeling task. More broadly, our results support the growing view that high-quality self-supervised representations can serve as reusable foundations for multiple downstream clinical applications, reducing the need to repeatedly train task-specific representation models for different datasets or monitoring conditions [5,11,12].

Beyond the methodological contributions, the public release of the SafeICU database together with pretrained PhysioStack models provides a valuable resource for the critical care research community. Although publicly accessible critical care databases such as MIMIC-III, eICU, and the PIC database have substantially accelerated the development and benchmarking of clinical AI, the availability of large-scale, high-frequency pediatric physiological datasets remains limited [10,17,18]. This limitation is particularly important because physiological responses, disease progression, and treatment strategies differ considerably between children and adults, making direct translation of adult-derived models challenging [4,19]. SafeICU complements these existing resources by providing longitudinal multimodal ICU data collected from a real-world pediatric critical care environment, including continuous physiological monitoring, laboratory investigations, treatment records, microbiology, and clinical outcomes. By releasing both the curated dataset and pretrained representation models, we aim to lower the barrier for developing, reproducing, and externally validating physiological foundation models while reducing the substantial computational cost associated with large-scale self-supervised pretraining. We anticipate that these resources will facilitate future research in transferable physiological representation learning, multimodal clinical foundation models, and AI development for pediatric and resource-constrained critical care settings [5,4,19,20].

The ability to transfer learned physiological representations across heterogeneous temporal resolutions has important implications for the clinical deployment of AI in critical care. In practice, intensive care units differ substantially in monitoring infrastructure, data acquisition systems, and clinical workflows, resulting in physiological data being recorded at varying sampling frequencies across institutions and healthcare settings [3,9,19]. Conventional machine learning models are typically developed and validated using data collected at a single temporal resolution, often requiring retraining or extensive recalibration before deployment in environments with different monitoring practices. Such requirements increase computational cost, limit scalability, and hinder widespread clinical adoption [3,9]. Our findings suggest that pretrained physiological representations can mitigate these challenges by providing a common representation space that remains informative across multiple temporal resolutions. This capability offers a practical pathway toward deploying a single pretrained representation model across diverse healthcare environments while allowing lightweight downstream models to be adapted for specific clinical applications. Such flexibility may be particularly valuable for pediatric and resource-constrained healthcare settings, where standardized high-frequency monitoring data are often unavailable but clinically robust AI decision-support tools are urgently needed [4,19].

This study has several limitations that provide opportunities for future research. First, although PhysioStack demonstrated robust temporal resolution transfer and cross-cohort generalization using independent pediatric and adult ICU cohorts, the representation model was pretrained on a single pediatric dataset. Evaluating large-scale self-supervised pretraining across multiple geographically and clinically diverse institutions will be important for further establishing the robustness and scalability of transferable physiological representations [6,9]. Calibration of the model was maintained more effectively within the source population than during transport to a distinct cohort (Figure 4, 5). In the cross cohort comparison, risk was overestimated among low-risk patients and underestimated among intermediate risk patients. This limitation may arise from differences in case mix, event prevalence, clinical practice, population characteristics or measurement processes, although the model is still well calibrated for high risk patients. Importantly, the similarity of the calibration curves from 5- to 60-minute resolutions in both settings suggests that this drift is driven primarily by cohort shift rather than temporal downsampling, indicating the successful demonstration of our primary hypothesis around resolution transfer. Overall, we recommend that the probability estimates should not be assumed to transfer unchanged across institutions; local recalibration, and prospective verification of calibration intercept, slope and clinical thresholds, may therefore be required before deployment. Second, the current framework was developed using four routinely monitored physiological signals. Incorporating additional data modalities, including laboratory investigations, medications, clinical notes, and imaging, may enable richer multimodal representations capable of supporting a broader range of clinical decision-support tasks [5,13]. Third, although early shock prediction provides a clinically relevant validation of representation quality, future work should evaluate the pretrained representations across additional downstream applications such as sepsis, respiratory deterioration, mortality prediction, and length-of-stay estimation to better characterize their general utility [3,11].

## Conclusion and future directions

The findings of this study have direct relevance for the deployment of data-driven decision-support tools in intensive care. In routine clinical environments, particularly in low- and middle-income countries, continuous high-frequency monitoring is often unavailable, and physiological data are frequently documented at irregular or coarser temporal resolutions. Physiological representations pretrained on high-resolution data remained stable when transferred to lower monitoring frequencies. This finding suggests that models developed in well-resourced settings may be deployed across hospitals with heterogeneous monitoring practices without requiring resolution-specific retraining.

From a clinical systems perspective, resolution-robust representations may reduce implementation barriers for early warning and risk stratification tools by enabling a shared pretrained embedding backbone to support multiple downstream applications. This approach could improve consistency of model behavior across different settings and reduce the need for repeated local redevelopment when monitoring infrastructure differs. The released pretrained models provide a reusable foundation for future research and deployment, enabling the extraction of clinically meaningful physiological representations from new datasets without extensive retraining. Although shock prediction was used here only as an illustrative downstream task, the preservation of discrimination across temporal resolutions supports the broader hypothesis that representation-level robustness can improve reliability under real-world data constraints.

Future work should focus on prospective evaluation in additional pediatric ICUs with differing monitoring infrastructure and case mix to establish clinical usability and external validity. Extending this framework to multimodal ICU data, including laboratory measurements, medications, and clinical notes, will be important to determine whether resolution-robust pretraining can support more comprehensive decision-support systems. Finally, integration of such representations into clinician-facing workflows will require careful evaluation of interpretability, calibration, and safety to ensure alignment with bedside decision-making.

## Data Availability

The SafeICU pediatric intensive care unit database released as part of this study is available at https://safeicu.aiims.edu.in
. Access to the dataset is available to credentialed users upon completion of a data use agreement, subject to institutional and ethical requirements. Pretrained PhysioStack representation models, together with code for data preprocessing, model training, and evaluation, are publicly available at https://github.com/tavlab-iiitd/PhysioStack
.

https://safeicu.aiims.edu.in

## List of Abbreviations

ABP: Arterial Blood Pressure
AIIMS: All India Institute of Medical Sciences
HIPAA: Health Insurance Portability and Accountability Act
HMAC: Hash-based Message Authentication Code
HR: Heart Rate
ICUs: Intensive Care Units
MIMIC: Medical Information Mart for Intensive Care
MLM: Masked Language Model
PBICDF2: Password-Based Key Derivation Function 2
PICU: Pediatric Intensive Care Unit
RECORD: REporting of studies Conducted using Observational Routinely-collected health Data
RR: Respiratory Rate
SHA256: Secure Hash Algorithm256
SI: Shock Index
SpO_2_: Oxygen Saturation
STROBE: Strengthening The Reporting of Observational studies in Epidemiology

## Declarations

### i) Funding

This research work was supported by the SafeICU Initiative, funded by the Indian Council of Medical Research (ICMR) under grant award number A1-Ad-hoc/18/2022-A1-Cell; the Centre of Excellence in Healthcare (COEH) at IIIT-Delhi; and Wellcome Trust/DBT India Alliance Fellowship (IA/CPHE/14/1/501504) awarded to Prof Tavpritesh Sethi.

### ii) Conflicts of interest

The authors declare that they have no competing interests

### iii) Ethics approval

- The study received ethical approval from the Ethics Committee of AIIMS Delhi under the reference numbers IEC/NP-211/08·05·2015 and IEC-787/07.10.2022, RP-14/2022.
- This observational study was conducted and reported in accordance with the STROBE (Strengthening the Reporting of Observational Studies in Epidemiology) guidelines.
- As the data used were routinely collected during standard clinical care, reporting was also guided by the RECORD (REporting of studies Conducted using Observational Routinely-collected health Data) statement.

### iv) Consent to participate

- Ethics approval was obtained for this study, and the requirement for individual consent was waived because all data were anonymized before use.

### v) Consent for publication

Not applicable

### vi) Code availability

The code underlying this study is available at https://github.com/tavlab-iiitd/PhysioStack

### vii) Availability of data and materials

The SafeICU pediatric intensive care unit database is released as part of this study and is available at safeicu.aiims.edu.in. Access to the dataset is granted to credentialed users following completion of a data use agreement (DUA), in accordance with institutional and ethical requirements.

Pretrained PhysioStack representation models, along with code for data preprocessing, model training, and evaluation, are publicly available at https://github.com/tavlab-iiitd/PhysioStack. Detailed documentation and usage instructions are provided in the accompanying README files to facilitate reproducibility and reuse.

### viii) Authors’ contributions

TS, RL, AD, PS, and JS conceived the study and defined the overall research objectives. PS, AD, TS, and RL managed data curation. AD and PS conducted the formal analysis and visualized the results. AD and PS led the data preprocessing, representation learning experiments, temporal resolution analyses, and downstream evaluations, and performed all statistical analyses. TS, JS, and RL supervised the study, guided the methodological design and interpretation of results, and contributed to manuscript writing and critical revision for intellectual content. AD and PS supported the SafeICU data release by developing and maintaining the data publication backend, managing server infrastructure, handling access control and credentialing workflows, and assisting with deployment, storage, and IP management required for controlled data sharing. The first draft of the report was written by AD and PS, with critical revisions made for important intellectual content by TS, RL, AD, and PS. All authors read and approved the final version of the manuscript and agree to be accountable for all aspects of the work.

## ix) Acknowledgments

The authors acknowledge funding support from the Indian Council of Medical Research (ICMR; AI-Adhoc-18/2022-AI Cell), the Wellcome Trust/DBT India Alliance Fellowship (IA/CPHE/14/1/501504) awarded to Dr. Tavpritesh Sethi, and the Centre of Excellence in Healthcare (COEH), IIIT-Delhi. The authors also thank the Department of Computational Biology, IIIT-Delhi, for infrastructural support. We sincerely thank the D5-ICU/MCB-PICU team for their technical assistance, and Anil Sharma and Varun Prakash at the Pediatric Intensive Care Unit of AIIMS, New Delhi. We also acknowledge the AIIMS computer facility for providing the computational infrastructure and secure access required for data storage, processing, and analysis. We specially thank Ranjeet Singh (AIIMS, New Delhi IT) and Bhawani Shah (IIIT-Delhi IT) for their support in establishing the network infrastructure required for SafeICU database development. Finally, we acknowledge Deepankar Verma, Likhith Devadiga, Pawan Yadav, and Shivam Mishra for their assistance with database vulnerability testing and security clearance.

## Notes

### Competing Interest Statement

The authors have declared no competing interest.

### Author Declarations

The Ethics Committee of the All India Institute of Medical Sciences, New Delhi, gave ethical approval for this work. The requirement for individual informed consent was waived because this study involved retrospective analysis of routinely collected anonymized clinical data.

### Summary of Updates

Added a new section on cross-cohort generalization evaluating downstream early shock prediction using transferred PhysioStack representations on an independent adult eICU cohort. Included new performance results across multiple temporal resolutions (Table 3), new ROC, precision-recall, and calibration analyses (Figure 5), and accompanying discussion demonstrating the transferability and robustness of the learned physiological representations.

## References

[1] Alsabri, M., Mourid, M.R., Saleh, A.R. et al. Vital signs as biomarkers of early clinical deterioration in pediatric emergency departments: physiology, interpretation, and innovations: a narrative review. Int J Emerg Med 19, 6 (2026). 10.1186/s12245-025-01107-8.

[2] Mayampurath, Anoop PhD1,2; Jani, Priti MD, MPH1; Dai, Yangyang MA2; Gibbons, Robert PhD3; Edelson, Dana MD, MS3; Churpek, Matthew M. MD, MPH, PhD3. A Vital Sign-Based Model to Predict Clinical Deterioration in Hospitalized Children*. Pediatric Critical Care Medicine 21(9):p 820–826, September 2020. | DOI: 10.1097/PCC.0000000000002414.

[3] Hong N., Liu C., Gao J., et al. State of the Art of Machine Learning–Enabled Clinical Decision Support in Intensive Care Units: Literature Review. JMIR Med Inform 2022;10(3):e28781, DOI: 10.2196/28781.

[4] Ganatra HA. Machine Learning in Pediatric Healthcare: Current Trends, Challenges, and Future Directions. J Clin Med. 2025 Jan 26;14(3):807. doi: 10.3390/jcm14030807. PMID: 39941476; PMCID: PMC11818243.

[5] Moor M., Banerjee O., Abad Z.S.H. et al. Foundation models for generalist medical artificial intelligence. Nature 616, 259–265 (2023). 10.1038/s41586-023-05881-4.

[6] Röösli E., Bozkurt S., Hernandez-Boussard T. Peeking into a black box, the fairness and generalizability of a MIMIC-III benchmarking model. Sci Data 9, 24 (2022). 10.1038/s41597-021-01110-7.

[7] Zhang K., Fan Y., Long K., Lan Y., Gao P. Research Hotspots and Trends of Deep Learning in Critical Care Medicine: A Bibliometric and Visualized Study. J Multidiscip Healthc. 2023;16:2155–2166. 10.2147/JMDH.S420709.

[8] Zech JR, Badgeley MA, Liu M, Costa AB, Titano JJ, et al. (2018) Variable generalization performance of a deep learning model to detect pneumonia in chest radiographs: A cross-sectional study. PLOS Medicine 15(11): e1002683. 10.1371/journal.pmed.1002683

[9] Kelly, C.J., Karthikesalingam, A., Suleyman, M. et al. Key challenges for delivering clinical impact with artificial intelligence. BMC Med 17, 195 (2019). 10.1186/s12916-019-1426-2

[10] Pollard T., Johnson A., Raffa J. et al. The eICU Collaborative Research Database, a freely available multi-center database for critical care research. Sci Data 5, 180178 (2018). 10.1038/sdata.2018.178.

[11] Liu Z., Alavi A., Li M., Zhang X. Self-Supervised Contrastive Learning for Medical Time Series: A Systematic Review. Sensors. 2023; 23(9):4221. 10.3390/s23094221

[12] Li, Y., Rao, S., Solares, J.R.A. et al. BEHRT: Transformer for Electronic Health Records. Sci Rep 10, 7155 (2020). 10.1038/s41598-020-62922-y

[13] Yang, X., Chen, A., PourNejatian, N. et al. A large language model for electronic health records. npj Digit. Med. 5, 194 (2022). 10.1038/s41746-022-00742-2

[14] Devlin J., Chang M., Lee K., Toutanova K. (2019). BERT: Pre-training of Deep Bidirectional Transformers for Language Understanding. North American Chapter of the Association for Computational Linguistics.

[15] Brown TB., Mann B., Ryder N, et al. 2020. Language models are few-shot learners. In Proceedings of the 34th International Conference on Neural Information Processing Systems (NIPS ’20). Curran Associates Inc., Red Hook, NY, USA, Article 159, 1877–1901.

[16] Bommasani, Rishi et al. “On the Opportunities and Risks of Foundation Models.” ArXiv abs/2108.07258 (2021): n. Pag

[17] Zeng X., Yu G., Lu Y. et al. PIC, a paediatric-specific intensive care database. Sci Data 7, 14 (2020). 10.1038/s41597-020-0355-4.

[18] Johnson A., Pollard T., Shen L. et al. MIMIC-III, a freely accessible critical care database. Sci Data 3, 160035 (2016). 10.1038/sdata.2016.35.

[19] Olatunji G., Kokori E., Aderinto N., et al. Challenges and Strategies in Pediatric Critical Care: Insights From Low-Resource Settings. Global Pediatric Health. 2024;11. doi:10.1177/2333794X241285964.

[20] Declerck J., Lee J., Sen A., et al. The Potential to Leverage Real-World Data for Pediatric Clinical Trials: A Proof-of-Concept Study. J Med Internet Res 2025;27:e72573. DOI: 10.2196/72573

[21] von Elm E., Altman DG., Egger M., Pocock SJ., Gøtzsche PC., Vandenbroucke JP. STROBE Initiative. The Strengthening the Reporting of Observational Studies in Epidemiology (STROBE) statement: guidelines for reporting observational studies. Lancet. 2007 Oct 20;370(9596):1453–7. doi: 10.1016/S0140-6736(07)61602-X. PMID: 18064739.

[22] Benchimol EI., Smeeth L., Guttmann A., Harron K., Moher D, Petersen I, Sørensen HT, von Elm E, Langan SM; RECORD Working Committee. The REporting of studies Conducted using Observational Routinely-collected health Data (RECORD) statement. PLoS Med. 2015 Oct 6;12(10):e1001885. doi: 10.1371/journal.pmed.1001885. PMID: 26440803; PMCID: PMC4595218.

[23] Atchinson BK., Fox DM. The politics of the Health Insurance Portability and Accountability Act. Health Aff (Millwood). 1997 May-Jun;16(3):146–50. doi: 10.1377/hlthaff.16.3.146. PMID: 9141331.

[24] Koch E, Lovett S, Nghiem T, Riggs RA, Rech MA. Shock index in the emergency department: utility and limitations. Open Access Emerg Med OAEM. 2019;11:179–99.

[25] Cheng TH, Sie YD, Hsu KH, Goh ZNL, Chien CY, Chen HY, et al. Shock Index: A Simple and Effective Clinical Adjunct in Predicting 60-Day Mortality in Advanced Cancer Patients at the Emergency Department. Int J Environ Res Public Health. 2020 Jul;17(13):4904.

[26] Birkhahn RH, Gaeta TJ, Tloczkowski J, Terry D, Bove JJ. The shock index in early acute hypovolemia. Acad Emerg Med. 2003;10(5):494–5.

[27] Stein, Dan Fredman, et al. Predicting Cardiovascular deterioration in a paediatric intensive care unit (PicEWS): a machine learning modelling study of routinely collected health-care data. eClinicalMedicine, Volume 85, 103255.

