## Supplementary Table for "Generalizing intensive care AI across time scales in resource-limited settings"

### Supplementary Material

**Supplementary Material 1.** Characteristics of PICU Admissions Included in the SafeICU Database. Summarizes demographic characteristics, length of stay, and outcomes for pediatric intensive care unit admissions included in the SafeICU database between January 2015 and December 2024.

| | **ICU admissions** |
|---|---|
| **Duration** | Jan 2015 - Dec 2024 |
| **Unique patients** | 2,275 |
| **Age (years)** | |
| Median (Q1-Q3) | 6.67 (3.5-9.9) |
| **Gender, n (%)** | |
| Male, n (%) | 651 (60.68%) |
| Female, n (%) | 422 (39.32%) |
| **ICU length of stay (days)** | |
| median (Q1-Q3) | 13 (6-23.5) |

**Supplementary Material 2.** Summarizes key characteristics of the SafeICU pediatric intensive care unit database in comparison with widely used critical care databases, including MIMIC-IV, the eICU Collaborative Research Database, and the Pediatric Intensive Care (PIC) database.

| | **MIMIC-IV** | **eICU** | **PIC** | **SafeICU** |
|---|---|---|---|---|
| **Language** | English | English | English-Chinese bilingual | English |
| **Data source** | 673-bed general hospital | Multi-center ICU | 1900-bed | 8-bed |
| **Category of critical care** | CCU, CSRU, MICU, SICU, TSICU, NICU. | eICU-CRD | CICU, SICU, PICU, NICU, GICU. | PICU |
| **Number of records** | 50,000+ | 200,000+ | 10,000+ | 2,000+ |
| **Patient age, median [IQR]** | 60.6 years (42.1-74.1) | 65 years (53-76) | 0.8 years (0.1-3.5) | 6.67 years (3.5-9.9) |
| **Vital signs** | Bedside monitor generated (1 minute) | Originally collected at 1-minute intervals | The daily nurse recorded and the surgery monitor (5 minutes) | Bedside monitor (15 seconds) |
| **Laboratory/clinical data** | Yes | Yes | Yes | Yes |
| **Clinician notes** | Yes | Yes | Extracted symptoms from notes | Nurse-documented treatment notes |
| **Mortality** | Death recorded in the hospital and Social Security database | Death recorded in the hospital | Death recorded in the hospital | Death recorded in the hospital |

*Abbreviations: CCU, coronary care unit; CICU, cardiac intensive care unit; CSRU, cardiac surgery recovery unit; GICU, general intensive care unit; ICU, intensive care unit; MICU, medical intensive care unit; NICU, neonatal intensive care unit; PICU, pediatric intensive care unit; SICU, surgical intensive care unit; TSICU, trauma/surgical intensive care unit.*

**Supplementary Material 3.** Hyperparameter Optimization for Representation Learning.

To identify optimal training configurations for representation learning, automated hyperparameter optimization was performed using Optuna, a Bayesian optimization framework. The optimization objective was to maximize MLM accuracy on a held-out validation set.

Each optimization trial involved training a transformer-based model with a unique combination of hyperparameters sampled from predefined search spaces. The hyperparameters explored included optimizer type, learning rate, batch size, weight decay, and learning rate scheduler. Optimization trials were parallelized across available graphics processing units to accelerate convergence. To limit unnecessary computation, early stopping was applied, and training was terminated if validation performance did not improve for 25 consecutive epochs.

Hyperparameter optimization was performed independently for each physiological signal and temporal resolution. The resulting optimized configurations were used for all representation learning experiments reported in the main manuscript unless otherwise specified.

| **Hyperparameter** | **Search Space** |
|---|---|
| Optimizer | Adam, SGD, RMSprop |
| Learning rate | $3 \times 10^{-5}$ to $3 \times 10^{-3}$ |
| Scheduler | Linear, Cosine |
| Batch size | 4, 8, 16, 32 |
| Weight decay | $1 \times 10^{-5}$ to $1 \times 10^{-3}$ |

---

**Supplementary Material 4.** Cohort Construction

Data from the Electronic ICU (eICU) database and the SafeICU pediatric database were used to construct the study cohorts. The eICU cohort included patients admitted to multiple ICU types, including medical (MICU), surgical (SICU), medical-surgical ICU, coronary care (CCU), cardiac ICU, cardiac surgery ICU (CSICU), cardiothoracic ICU (CTICU), and neuro-ICU units.

For both datasets, inclusion criteria required continuous vital sign monitoring for at least 7.5 hours during the ICU stay. Exclusion criteria were applied consistently and included: (i) absence of invasive arterial blood pressure measurements during the observation window; (ii) missing demographic information (age or sex), or age greater than 89 years where applicable; (iii) evidence of shock within the initial observation period to avoid bias from patients already exhibiting the outcome of interest; and (iv) missing data exceeding 10% of the observation window.

Since SafeICU is a pediatric data resource, different SI thresholds were used for various age groups (SI >2.3 (ages <= 3 months), >1.7 (4-6 months), >1.5 (7-12 months), >1.2 (13-36 months), >1.15 (37-72 months), >0.95 (73-144 months), >0.77 (>144 months))(22).

---

**Supplementary Material 5.** Representation Learning Performance Across Temporal Resolutions in the Adult ICU Cohort.

Summarizes MLM performance across physiological signals and temporal resolutions in the combined adult ICU cohort derived from the MIMIC-III and eICU databases. Median performance (interquartile range) is reported for HR, RR, $SpO_2$, and ABP. Results at 5-minute resolution are shown for both baseline and hyperparameter-optimized models; results at all other resolutions reflect optimized models.

| Temporal resolution | Training configuration | MLM accuracy, HR(Median [IQR]) | MLM accuracy, RR(Median [IQR]) | MLM accuracy, $SpO_2$(Median [IQR]) | MLM accuracy, ABP(Median [IQR]) |
|---|---|---|---|---|---|
| 5minutes | Baseline hyperparameters | 0.67 (0.24) | 0.45 (0.3) | 0.45 (0.3) | 0.38 (0.23) |
| 5minutes | Optimized hyperparameters | 0.87 (0.18) | 0.74 (0.23) | 0.79 (0.21) | 0.79 (0.21) |
| 10minutes | Optimized hyperparameters | 0.83 (0.18) | 0.68 (0.26) | 0.73 (0.2) | 0.71 (0.21) |
| 15minutes | Optimized hyperparameters | 0.79 (0.21) | 0.64 (0.29) | 0.7 (0.24) | 0.64 (0.22) |
| 30minutes | Optimized hyperparameters | 0.74 (0.29) | 0.60 (0.27) | 0.61 (0.3) | 0.5 (0.26) |
| 60minutes | Optimized hyperparameters | 0.57 (0.42) | 0.56 (0.42) | 0.56 (0.41) | 0.2 (0.33) |

*Abbreviations: ABP, arterial blood pressure; HR, heart rate; IQR, interquartile range; MLM, masked language modeling; RR, respiratory rate; $SpO_2$, oxygen saturation.*

---

**Supplementary Material 6.** Shock Prediction Model Selection and resolution transfer on adult data.

#### Shock Classification Models

Four standard classification models were evaluated for early shock prediction using physiological embeddings as inputs:

- Gradient Boosting Classifier (GBC)
- Support Vector Machine (SVM)
- Random Forest (RF)
- Logistic Regression (LR)

All models were trained using embeddings extracted from pretrained language models and evaluated under consistent data splits, as described in the main Methods.
Class Imbalance Handling. For all models, class imbalance in the training set was addressed using random undersampling to achieve a 1:1 ratio of shock and non-shock samples. Undersampling was applied only to training data and not to validation or test sets.

#### Five-Fold Cross-Validation Performance for Shock Prediction Models

Summarizes five-fold cross-validation performance of shock prediction models evaluated using physiological embeddings derived from pretrained language models. Median performance and interquartile range are reported across folds. Embeddings were generated from 5-minute resolution data using resolution-matched pretrained language models.

| Model | Accuracy, Median (IQR) | Precision, Median (IQR) | Recall, Median (IQR) | F1 Score, Median (IQR) | AUC, Median (IQR) | AUPRC, Median (IQR) |
|---|---|---|---|---|---|---|
| Gradient Boosting | 0.82 ± 0.03 | 0.82 ± 0.04 | 0.95 ± 0.02 | 0.88 ± 0.02 | 0.87 ± 0.01 | 0.92 ± 0.01 |
| Random Forest | 0.78 ± 0.01 | 0.80 ± 0.01 | 0.91 ± 0.03 | 0.85 ± 0.02 | 0.83 ± 0.01 | 0.90 ± 0.01 |
| Logistic Regression | 0.68 ± 0.03 | 0.68 ± 0.03 | 0.99 ± 0.01 | 0.81 ± 0.03 | 0.64 ± 0.01 | 0.77 ± 0.04 |
| SVM | 0.68 ± 0.03 | 0.68 ± 0.03 | 0.99 ± 0.00 | 0.81 ± 0.02 | 0.62 ± 0.03 | 0.75 ± 0.04 |
